# Bridging and analytical validation of the Prosigna^®^ Breast Risk of Recurrence Test as a whole-transcriptome NGS lab developed test

**DOI:** 10.64898/2026.06.23.26355479

**Authors:** Dongyang Zhang, Ying Wang, Lindsay Sager, Rachel Koenigsberg, Mugdha Birari, Alex Hakansson, Emily Fogarty, Jason W. Reeves, Carlo Artieri, Lori Lofaro, Hege G. Russnes, Hege O. Ohnstad, Bjørn Naume, Phillip G. Febbo, Paul K. Marcom, Jeff Gole

## Abstract

**Background:** The Prosigna Breast Risk of Recurrence test is based on the PAM50 classifier and was originally validated as an *in vitro* diagnostic (IVD) test on the Dx enabled nCounter^®^ Analysis System. The Prosigna test is intended for early-stage, hormone receptor+ (HR+) breast cancer and provides the risk of recurrence (ROR) score (0-100), intrinsic subtype (Luminal A, Luminal B, HER2-enriched, and Basal-like), and the 10-year probability of distant recurrence. We describe the performance of the Prosigna test as a whole transcriptome RNA sequencing laboratory developed test (LDT) for measuring the Prosigna ROR score and intrinsic subtypes on tissue from surgical resection and core needle biopsy as compared to the Prosigna test on the nCounter system.

**Methods:** We evaluated three separate breast cancer cohorts to 1) bridge the IVD test on the nCounter system and NGS LDT test (n = 245), 2) validate the bridged algorithm on an independent biobank sample set (n = 187), and 3) retrospectively test performance on long-term archival samples from a previous study (n = 109).

**Results:** Bridging analysis showed minimal score variability and robust correlation of Prosigna ROR scoring in surgical resections (SR) (2.459, SD; 0.981, R^2^) and core needle biopsy (CNB) (2.338, SD; 0.970, R^2^) samples. In the validation set, the Prosigna NGS LDT ROR scores maintained high correlation to the scores of the nCounter system (SR = 0.968, CNB = 0.966, R^2^), exhibited minimal score variability (SR = 2.488, CNB = 2.558, SD), and demonstrated high concordance in subtype classifications (SR = 92.3% CNB = 92.8%). Further testing demonstrated comparable performance across tumor fractions, a lower limit of detection (LLOD) of 5 ng, and robustness to exogenous ethanol or genomic DNA contamination. When testing previously extracted RNA from the clinical cohort, we observed high correlation (0.974, R^2^) and low variance (3.078, SD) of ROR scores with original values on the nCounter system, along with strong risk group (95.4%) and subtype (94.5%) concordance.

**Conclusions:** This study describes the analytical validation of the Prosigna NGS-based LDT measuring the Prosigna ROR score and intrinsic subtypes with robust analytical performance on SR and CNB specimens, providing confidence for clinicians utilizing the NGS-based version of this well-established test.

## Background

Gene expression profiling (GEP) tests have become a backbone for assessing individualized risk for breast cancer patients over the last two decades (1–10). The Prosigna Breast Risk of Recurrence test is a GEP test that is based on the PAM50 classifier and combines intrinsic subtypes (Luminal A, Luminal B, Basal-like, and HER2-enriched) (11, 12), tumor proliferation, and tumor size to calculate a patient’s risk of recurrence (ROR) score (1, 2, 4, 5, 13, 14). The ROR score is used to estimate a probability of distant recurrence at 10 years, based on a patient receiving 5 years of endocrine therapy.

Ongoing prospective randomized phase 3 trials are evaluating test-directed prediction of chemotherapy benefit with the Prosigna test in high-risk HR+ early-stage breast cancer (EBC) patients (OPTIMA, ISRCTN42400492) (15) and younger premenopausal HR+ EBC patients (OPTIMA-YOUNG, NCT07106632) (16), and prediction of response to radiation treatment in low-risk EBC (EXPERT, NCT02889874) (17). While those trials are still active, the prognostic utility of the Prosigna ROR score has been well established (3, 5, 13, 14, 18, 19). Moreover, analysis of the Prosigna test and research surrogates of the test, have found that the Prosigna ROR score is an accurate prognostic risk of recurrence model in diverse populations (20), and can provide accurate long-term 10-year distant recurrence prognosis (13, 14, 21).

In addition to the Prosigna ROR score, the ability molecularly classify breast tumor subtype has been recognized as a critical advancement for researching and treating breast cancer (22, 23). The Cancer Genome Atlas (TCGA) validated the ability of the intrinsic subtypes to capture the general molecular landscape of breast cancer across comprehensive molecular profiling techniques (24). Additionally, the PAM50 intrinsic subtypes have consistently been shown to be complementary to standard immunohistochemistry (IHC) testing for breast cancer (19, 25–28) and to be prognostic and predictive of patient outcomes with endocrine therapy (29, 30) and chemotherapy (31–35). While other subtype assignment methods have been developed (36), the concordance between tests is modest (37).

Previous studies have shown that the Prosigna test is applicable on both surgical resections (SR) as well as core needle biopsies (CNB) (1, 38, 39). Subsequent decision-impact studies have shown that clinicians benefit from having access to this test in both adjuvant and neoadjuvant settings when evaluating treatment options with patients (40–42). Therefore, providing access to this test has significant value as the landscape for treatment for breast cancer continues to evolve.

At the time that the Prosigna test was developed, few technologies existed that could produce a robust clinical test for gene expression signatures that were viable on routinely processed formalin-fixed paraffin embedded (FFPE) tissues. The nCounter Analysis System was selected for Prosigna test development due to the platform’s robust handling of degraded RNA, low RNA input requirements, and ability to multiplex more than 50 targets into a single assay (39, 43, 44). Since the original development, advances in RNA sequencing have closed the gap between the technologies, and sequencing extracted nucleic acid from FFPE has become routine in research settings. However, even with these advancements, prognostic and predictive GEP breast cancer diagnostics rely on targeted expression testing (6–8, 45). In contrast, research application of RNA-sequencing for FFPE often focuses on whole transcriptome analysis driven by broader exome capture panels (46–48). Several LDTs have been published that leverage transcriptome-wide RNA sequencing (49–51), but these have been developed to primarily address other clinical and translational research questions rather than serving as prognostic or predictive diagnostic GEP tests for breast cancer.

Herein we describe a multi-stage study to first bridge the original Prosigna test for the nCounter system with a newly developed NGS LDT and then validate the performance of the Prosigna NGS ROR score and intrinsic subtype classifications in two additional independent cohorts. This study was designed to validate the equivalence of the Prosigna test when run as a centrally tested LDT that takes advantage of the improved performance and scalability of NGS. An exome-capture whole transcriptome RNA sequencing platform was developed for calculating the patient ROR score and intrinsic subtype classification. As SR and CNB specimens support potential clinical intervention in the neoadjuvant and adjuvant setting respectively, we balanced our study to assess both specimen types and ensure we appropriately account for potential sources of pre-analytical variability from these distinct specimens.

## Methods

### Breast cancer samples and synthetic controls

FFPE blocks from surgical resection (SR, n = 222) and core needle biopsy (CNB, n = 210) samples were obtained from a commercial tissue bank (French Tissue Bank). Samples were allocated to two cohorts for bridging (n = 118 SR, 127 CNB) and analytical validation (n = 104 SR, 83 CNB). Additionally, long-term archival, previously extracted RNA (n = 112) and archival FFPE blocks (n = 28) were tested from a longitudinal breast cancer study (Oslo1) (19) to assess assay performance as an independent clinical cohort. Patient and sample characteristics for tumor bank and archival samples are documented in **Table 1**. Cell line derived controls, high risk Prosigna control (HPC) and low risk Prosigna control (LPC), were included during bridging and analytical validation. Synthetic genomic DNA (gDNA) contamination was prepared from extracted DNA from NA12878 (Coriell) and sheared to an average size of 400 bp by sonication (Covaris).

### Sample processing and RNA extraction

Serial 5 µm sections were cut from FFPE blocks and mounted to glass slides for both SR and CNB samples. Tumor areas were marked on HCE-stained slides to identify regions of interest (ROIs) for macrodissection. For validation samples, tumor fraction was estimated by pathologist evaluation when evaluable HCE was available. FFPE tissue sections were macrodissected and scraped from unstained slides as previously described (39, 44). Six unstained sequential slides were prepared for CNB samples, or 10 unstained sequential slides for SR samples. Each set of slides was split into two sets of 3 slides for CNB or 5 slides for SR and extracted in parallel for testing with the nCounter system or NGS methods described below. Replicate RNA extraction was performed on 10 samples to assess precision during validation. Two macrodissected slides per replicate were extracted, and extractions were performed in triplicate per patient block for each extraction batch. RNA extraction was evaluated on 5 x 5 µm and 3 x 10 µm slides from the same block on 10 additional samples.

Samples were extracted utilizing either the Veracyte FFPE RNA Extraction Kit (Veracyte, 550100) for the nCounter system or MagMax FFPE DNA/RNA Ultra (Thermo Fisher) using AutoLys M Tubes (Hamilton) on a Kingfisher Apex system (Thermo Fisher) for NGS testing. A negative extraction control (NEC) was collected in an empty AutoLys tube to evaluate contamination. Extracted RNA was quantified on a Lunatic instrument (Unchained Labs). PHC and PLC samples were fragmented using heat degradation and confirmed to have a DV 200 of 50-70% on a TapeStation D5000 (Agilent).

### Prosigna testing on the nCounter system

Samples processed for benchmarking with the nCounter system were processed using v5 of the Instructions for Use of the IVD assay. All samples were evaluated for standard QC metrics for the nCounter system, including a minimum reference sample mean housekeeping gene count of 3000. Samples passing QC were classified for PAM50 IS subtype and ROR score as previously described (39, 44).

### NGS library preparation and sequencing

Following extraction, automated library preparation was performed on Hamilton Star liquid handling instruments utilizing the Twist Whole Transcriptome Library Preparation Kit using 50 ng of RNA or the maximum sample volume possible (11 µl) if the concentration was insufficient. 200 ng or the maximum sample volume (17.5 µl) of second PCR product was used for sequencing hybridization, with molarity and library size (bp) measured using the TapeStation D5000 (Agilent Technologies). For each test, 200-cycle sequencing runs were captured on 10B flow cells (Illumina) on a NovaSeq X+ (Illumina).

During bridging and validation, each library prep plate contained a mixture of SR and CNB samples, control samples, and non-template controls (NTCs). Contamination during library preparation was evaluated by reviewing the PCR1 concentration for each of the NTCs (cutoff of 14 ng/µl).

### Alignment, NGS QC, and Prosigna test calculation

As described previously for other Veracyte LDTs (52, 53), sequencing BCL files were converted into FASTQ files and demultiplexed using the BCL convert software from Illumina (version 4.2.7), aligned using SentieonStar (version 202112.07), and quantified with HTSeq (version 2.0.2). Each sample was assessed for standard NGS sequencing quality control metrics using RNA-SeQC (version 2.4.2).

Gene counts were normalized using the variance stabilizing transformation (VST) tool from the R DESeq2 software (version 1.38.0). PCA-based batch-correction and platform calibration was performed to account for primary sources of assay variabilities (e.g. reagents, operators, instruments). The Prosigna test scores, including ROR and intrinsic subtype, were then calculated from the normalized NGS data for each sample using the previously described methods and model weightings (39, 44). In brief, this includes calculating the correlation of a sample to the intrinsic subtype centroids, calculating the tumor proliferation score, and weighting these values along with the tumor size to calculate a final ROR score as defined previously.

### Statistical Analysis

All statistical analysis was performed using Python (version 3.12) and R (version 4.4).

Predefined acceptance criteria were established based on simulations and previously established thresholds for the IVD assay. An acceptable threshold of the cross-platform (nCounter versus NGS) standard deviation (SD) ≤ 5 ROR units was set based on simulation of clinical data from ABCSG-8 (3, 4). A two one-sided test (TOST) procedure with bounds of ±3 ROR units was further used to test score equivalence. Previously established risk category thresholds were used for node negative (low [0, 40]; intermediate [41, 60]; high risk [61, 100]) and node positive samples (low [0, 40]; high [41, 100]) (3). Previous analysis of precision for the Prosigna assay for the nCounter system found an SD of 2.9 ROR units. SD thresholds reflect real-world performance and correspond to result in major risk group reclassification (low-risk [0-40] to high-risk [61-100] and vice versa) rate ≤ 5% for all samples. Low-(0–40) and high-risk group (41–100) concordance for node positive samples should exceed 80%, and PAM50 IS concordance for all samples should exceed 85%. Major risk category and intrinsic subtype concordance were evaluated using categorical concordance (Pairwise Concordance, PWC). Intra-run and Inter-run PWC were evaluated for all possible pairs of samples. Post-hoc analysis of subtype and across all risk group categories used unweighted and linearly weighted Cohen’s κ, respectively (54).

Acceptance criteria for precision studies were set to achieve parity with the previously validated IVD assay for the nCounter system (39, 44). A total standard deviation threshold of ≤ 4.3, computed by variance decomposition using linear mixed effect models was allowed based on simulations and previous acceptance criteria for the assay for the nCounter system(44).

For assessment of the lower limit of detection (LLOD) and interfering substance testing with artificial ethanol or genomic DNA contamination, an additional acceptability criterion required the 90% confidence interval beyond the mean difference in ROR scores was entirely contained within [-3, 3] ROR units (44). Analysis was performed on samples that passed sequencing QC, and post-hoc analysis was run on the whole cohort regardless of sequencing QC.

### Variance Decomposition Analysis

For precision studies, replicates of the same sample were analyzed for sources of both intra- and inter-run ROR score variability. Linear mixed effect models (LMM; R lme4, version 1.1.38) were used to evaluate sources of variation as components of total assay variability for ROR. Each biological sample was modeled as a fixed effect, while experimental sources of variation were modeled as random effects, such as reagent lot, instrument, and operator.

Inter-run variability was defined as the total variation observed both between plates and within plates, shown as the total SD computed by the linear mixed-effects models. Intra-run variability was defined as the within-plate variability, shown as the residual SD derived from the linear mixed-effects models. The models were defined as:

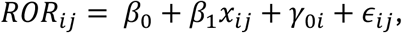

Where *β*_1_*x_ij_* represents the fixed effect, *γ*_0*i*_ represent the random intercept of the conditions and *ε_ij_* represents the residual error

## Results

### Study Design

To develop and evaluate the performance of a NGS LDT for the Prosigna test we evaluated 3 cohorts of breast cancer samples as well as control samples. These cohorts consisted of a platform bridging cohort (n = 245), an analytical validation cohort (n = 187), and a long-term archival real-world validation cohort (n = 112 RNA, 28 FFPE blocks) as shown in **Figure 1**. Sample characteristics of each cohort are described in **Table 1**. Samples were selected to represent a range of patient characteristics, ROR scores, and intrinsic subtypes where feasible. For the clinical cohort, samples were selected to be equally balanced over ROR and PAM50 IS classifications, with each subtype representing approximately a quarter of the samples profiled.

**Figure 1:**
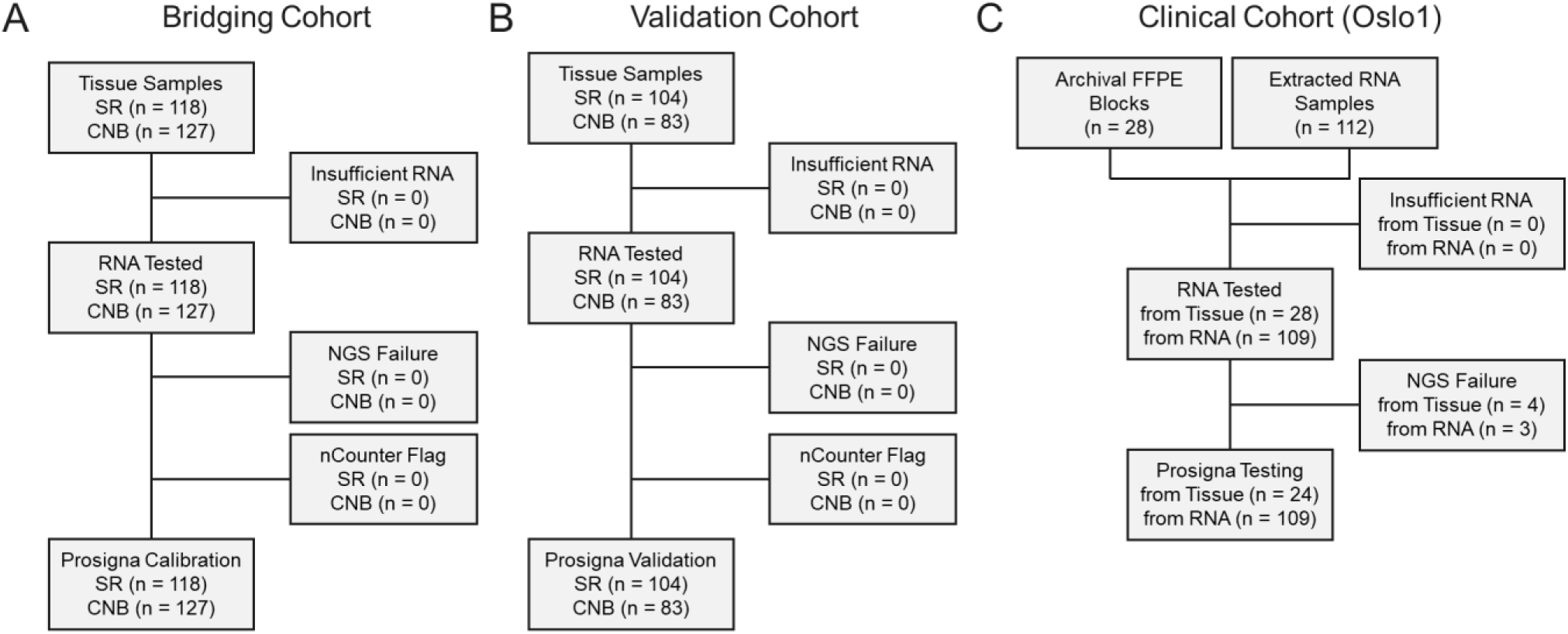
CONSORT diagram for bridging, validation and clinical cohorts. For bridging and validation cohorts the number of samples is shown for accuracy studies.

**Table 1:** Patient and sample characteristics of breast cancer specimens tested in each cohort. Unevaluable samples for tumor fraction did not have a paired H&E available prior to assessment.

|  | **Bridging Cohort** | **Validation Cohort** | **Oslo1 Cohort** |
| --- | --- | --- | --- |
| **Patient Samples** | 245 | 187 | 112 |
| Surgical Resection | 118 | 104 | 28 |
| Core Needle Biopsy | 127 | 83 | - |
| Archival RNA | - | - | 112 |
| **Patient Demographics** |  |  |  |
| *Age at Diagnosis* |  |  |  |
| Median | 43 | 66 | 59 |
| <55 | 167 | 68 | 41 |
| ≥55 | 78 | 119 | 71 |
| *Hormone Receptor Status* |  |  |  |
| HR+ | 225 | 170 | 109 |
| HR- | 20 | 17 | 3 |
| *Nodal Status* |  |  |  |
| N0 (0 nodes) | 137 | 121 | 67 |
| N1 (1-3 nodes) | 85 | 52 | 24 |
| N2+ (4+ nodes) | 23 | 14 | 18 |
| *Tumor Size* |  |  |  |
| 0-2 cm | 135 | 105 | 62 |
| >2 cm | 110 | 82 | 50 |
| **Specimen Characteristics** |  |  |  |
| *Tumor Fraction* |  |  |  |
| 10-40% | - | 11 | - |
| 50% | - | 13 | - |
| 60% | - | 15 | - |
| 70% | - | 56 | - |
| 80% | - | 62 | - |
| 90% | - | 21 | - |
| Not Evaluable | - | 9 | - |

### Bridging of the Prosigna test to NGS

To develop a platform bridging normalization algorithm for the NGS assay, a total of 245 patient specimens samples and controls, were tested across 11 plates, yielding 669 total replicates. As shown in **Supplementary Figure 1**, the bridging plates were sequenced using 3 reagent lots and 3 instruments across 3 sequencing runs with different laboratory operators.

Among the 11 total bridging plates, 4 pre-bridging plates were designed to screen the cohort and determine the impacts of platform, sample type, and experimental batch conditions. The pre-bridging plates contained only 1 replicate each per specimen sequenced. Principal component analysis (PCA) showed that specimen type was associated with 2 distinct clusters (**Supplementary Figure 2**). Therefore, eight samples (4 CNB, 4 SR) were selected as sequencing calibration samples and were replicated across the remaining 7 bridging plates to assess and correct for technical variability in the NGS assay to be used in modeling variance of sequencing for normalization development.

Within the 7 remaining bridging plates, all bridging RNA samples were processed, including precision and intra-run variance replicates. Different library preparation reagents, automation instruments, and laboratory operators were used across these seven plates to capture technical variability. In addition, 10 samples were run triplicate at input amounts of 20 ng, 50 ng, and 100 ng to further assess inter- and intra-run variability along with HPC and LPC control samples.

Analysis of the calibration replicate samples from the 7 bridging plates was used to develop a PCA-based batch correction algorithm to normalize technical variability between runs and minimize known sources of variability such as GC-content bias. As shown in **Supplementary Figure 2**, the algorithm effectively reduced technical variability of calibration replicate samples tested across multiple bridging plates. A linear mixed model was used to perform variance decomposition and to evaluate technical variability before and after batch correction. Analysis excluded the calibration replicate samples used for batch-correction algorithm training and only evaluated the remaining bridging samples as a test dataset. In these samples, the batch correction algorithm significantly reduced technical variation. Before batch correction the total SD was 2.49, and after batch correction the total SD was 1.07 (a 57% reduction).

The batch-corrected NGS data, together with the data from the nCounter system, were used to finalize the platform normalization algorithm by also accounting for gene-level differences in dynamic range prior to application of the Prosigna ROR score and intrinsic subtype calculation. Satisfactory normalization performance was observed, with a cross-platform SD of 2.459 for SR samples and 2.338 for CNB samples. Furthermore, regression analysis demonstrated that the normalized NGS assay produced consistent ROR scores compared to nCounter (SR = 0.981, CNB = 0.970, R^2^; SR = 0.995, CNB = 0.958 slope; **Figure 2B**). Similar results were observed after normalization for the IS centroid correlation (0.97 - 0.99, R^2^; 0.94 - 1.03, slope) and proliferation score (0.91 - 0.95, R^2^; 0.89 - 1, slope) components of ROR (**Supplementary Figure 3**).

**Figure 2:**
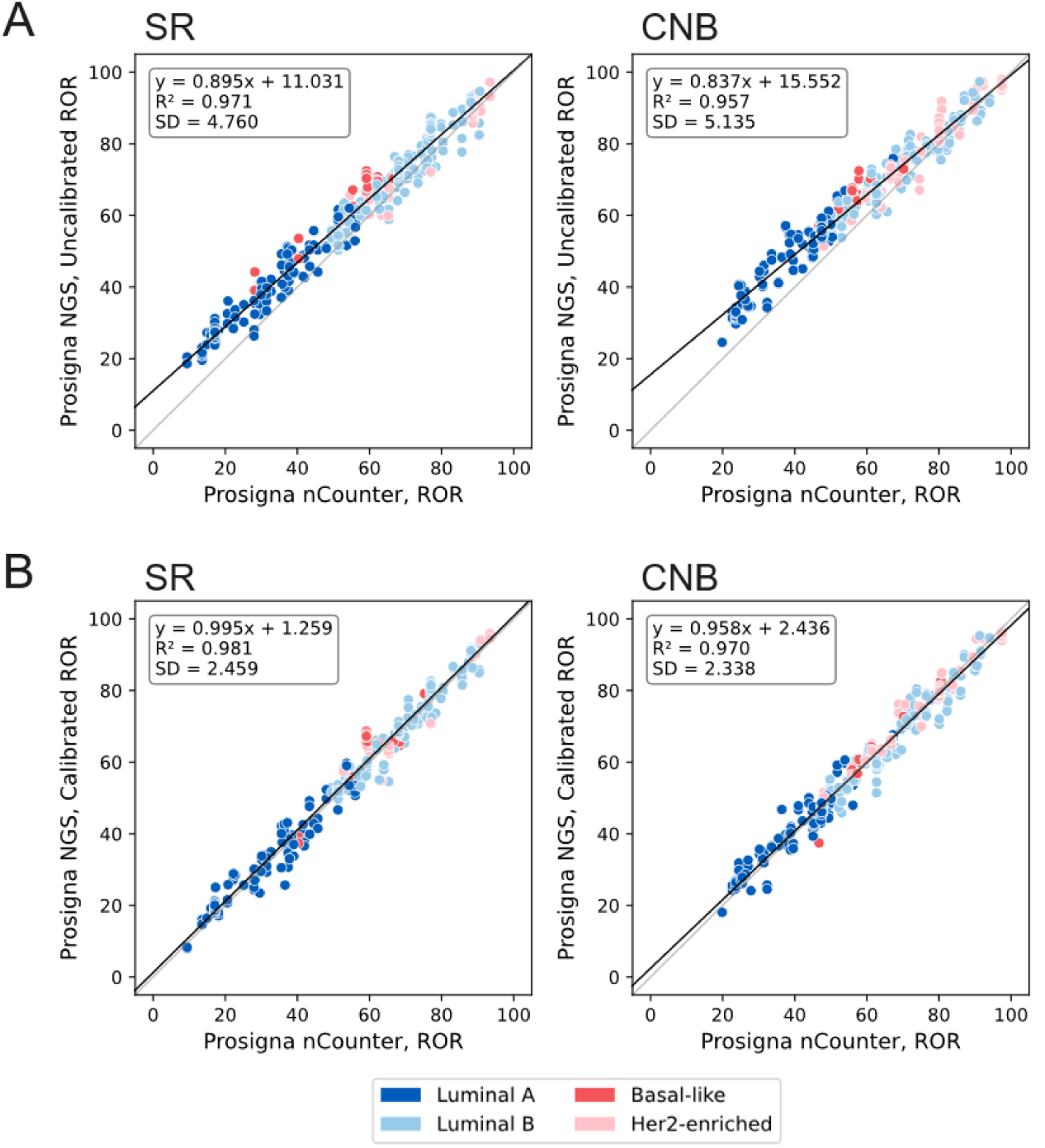
Linear regression of the Prosigna test ROR scores from bridging studies calculated from the nCounter system compared to A) raw VST gene expression from NGS or B) calibrated genes expression from NGS (n = 245). Analysis performed separately on SR (left) or CNB (right) samples. Samples are colored by intrinsic subtype from NGS analysis.

During bridging, intra-run and inter-run variability were assessed by testing selected samples in triplicate across multiple sequencing runs. Samples tested for precision were evaluated for sources of variation across 11 plates, 3 sequencing runs, and 3 instruments, and various RNA input amounts (20, 50, 100 ng). Variation in ROR scores by platform was determined to be low across the study (SD = 1.02, LMM) and attributable variance was negligible for most of the parameters tested. Minor variation was attributable to RNA input (SD contribution = 0.094, LMM).

### Analytical validation of the Prosigna test for NGS

Upon completion of the bridging algorithm development, a separate cohort of samples (n = 104 SR, 83 CNB) was analyzed to validate the analytical accuracy of the normalized NGS data and subsequent application of the Prosigna ROR scoring algorithm. Primary findings from the analytical validation study are summarized in **Supplementary Table 1**. During analysis of analytical validation only one reproducibility sample failed PCR1 amplification, and no samples were flagged during sequencing outside of interfering substance testing.

The variation observed between nCounter and NGS test results was within acceptable range for both SR (2.488, SD) and CNB (2.558, SD). Concordance of the Prosigna ROR scores was high in both SR (0.968, R^2^; 1.004, slope), and CNB (0.966, R^2^; 0.972, slope) samples (**Figure 3A**). Formal evaluation of platform equivalence of the tests between expression platforms used the two one-sided tests (TOST) procedure for equivalence bounds ±3 ROR units. SR and CNB specimens were tested separately and together across the entire cohort, and we found equivalent performance within the specified bounds for each comparison (p < 0.001) (**Supplementary Table 2**).

**Figure 3:**
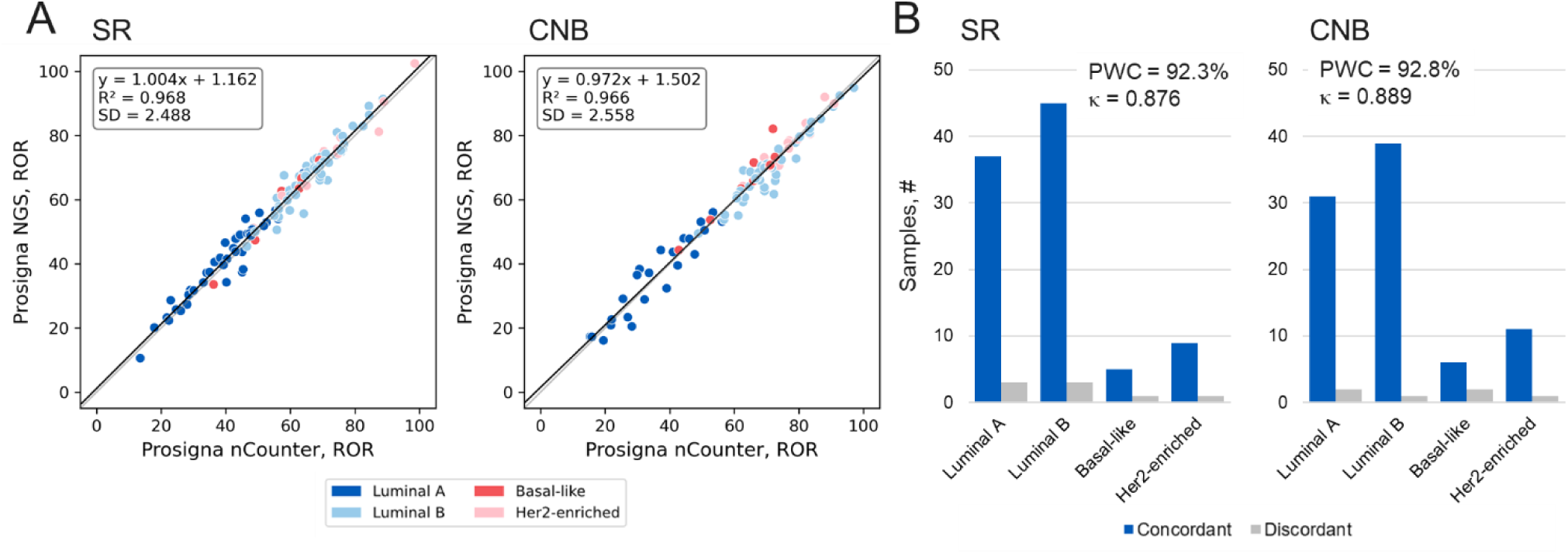
Concordance of the Prosigna test results from the nCounter system and NGS LDT results from the analytical validation study for ROR scores or intrinsic subtype classification (n = 189). A) Samples colored by intrinsic subtype classification from NGS. B) Samples with consistent intrinsic subtype classification colored in blue, discordant classification in gray. PWC and unweighted κ shown across all samples.

Samples were assigned major risk categories based on previously established thresholds across all patient samples (low risk ROR < 40, high risk ROR > 60) and specifically within samples from node positive patients (low risk ROR < 40, high risk ROR > 40). High pairwise concordance (PWC) was observed in major risk category analysis for all samples (SR = 100%, CNB = 100%, PWC) and node positive patient samples (SR = 97.67%, CNB = 95.45%). Post hoc analysis of concordance for all three risk categories found strong concordance in SR (87.50%, PWC; 0.846, weighted *κ*) and CNB (93.98%, PWC; 0.919, weighted *κ*) samples (**Supplementary Table 3**) (54). Similarly, we observed strong concordance of tumor intrinsic subtype in SR specimens (92.31%, PWC; 0.876, unweighted *κ*) and CNB specimens (92.77%, PWC; 0.889, unweighted *κ*) (**Figure 3B, Supplementary Table 4**) (54).

Analytical precision was tested by replicating extraction of RNA by varying experimental conditions (reagent lot, operator, instrument, n = 10 unique samples) and library preparation (reagent lot, operator instrument, n = 16 unique samples). Five independent library preparations were tested in triplicate at 50 ng input for a total of 15 technical replicates per biological sample. ROR score variability was acceptable for both RNA extraction (1.462, SD) and library preparation (0.951, SD). No significant variation was observed between sequencing runs (P > 0.5 ANOVA, **Supplementary Figure 4**). Furthermore, variance decomposition modeling did not identify any significant sampling variance attributable to reagent, instrument, and operator differences. Sequencing reproducibility was confirmed by evaluating 93 patient samples from the validation set on multiple sequencing runs and instruments at different read depths (10 billion and 1.5 billion reads for 93 patients, 1.5 billion reads for another subset of 36 patients). ROR risk and intrinsic subtype calls were consistent within these studies except for two samples (**Supplementary Table 5**), exceeding the acceptability criteria prespecified with >95% concordance among technical replicates in both evaluations.

To further qualify the assay, we evaluated whether the ROR score varied significantly across sample input specifications and with artificial contamination from interfering substances (**Figure 4, Supplementary Table 6**). To understand sample input parameters, we evaluated pathologist assessment of tumor fraction as well as RNA input requirements. Across the cohort, the ROR score performed within acceptance ranges (ROR SD ≤ 5) when compared across pathologist defined tumor fraction down to 10-40% tumor fraction (**Figure 4A**). A subset of 12 samples across ROR range and subtypes were selected from either bridging or validation samples to assess assay performance based on RNA input amount used to build NGS libraries. Samples were run in triplicate at an input of 2, 5, 10, 20, and 100 ng and compared to the 50 ng input ROR scores. All RNA inputs were observed to have acceptable variation (ROR SD ≤ 5), but 5 ng was the lowest input to meet secondary acceptance criteria (mean difference +/- 90% CI that was within ±3 ROR units) (**Figure 4B-C**). 2 ng input flags frequently failed NGS QC (23/36 samples, 63.9%), while no other input levels resulted in NGS QC flags during testing (**Figure 4C, Supplementary Table 6**). Additional extraction testing demonstrated that using a set of five 5 µm or three 10 µm slides both produced sufficient RNA to meet the 50 ng total input recommendation (**Supplementary Table 7**).

**Figure 4:**
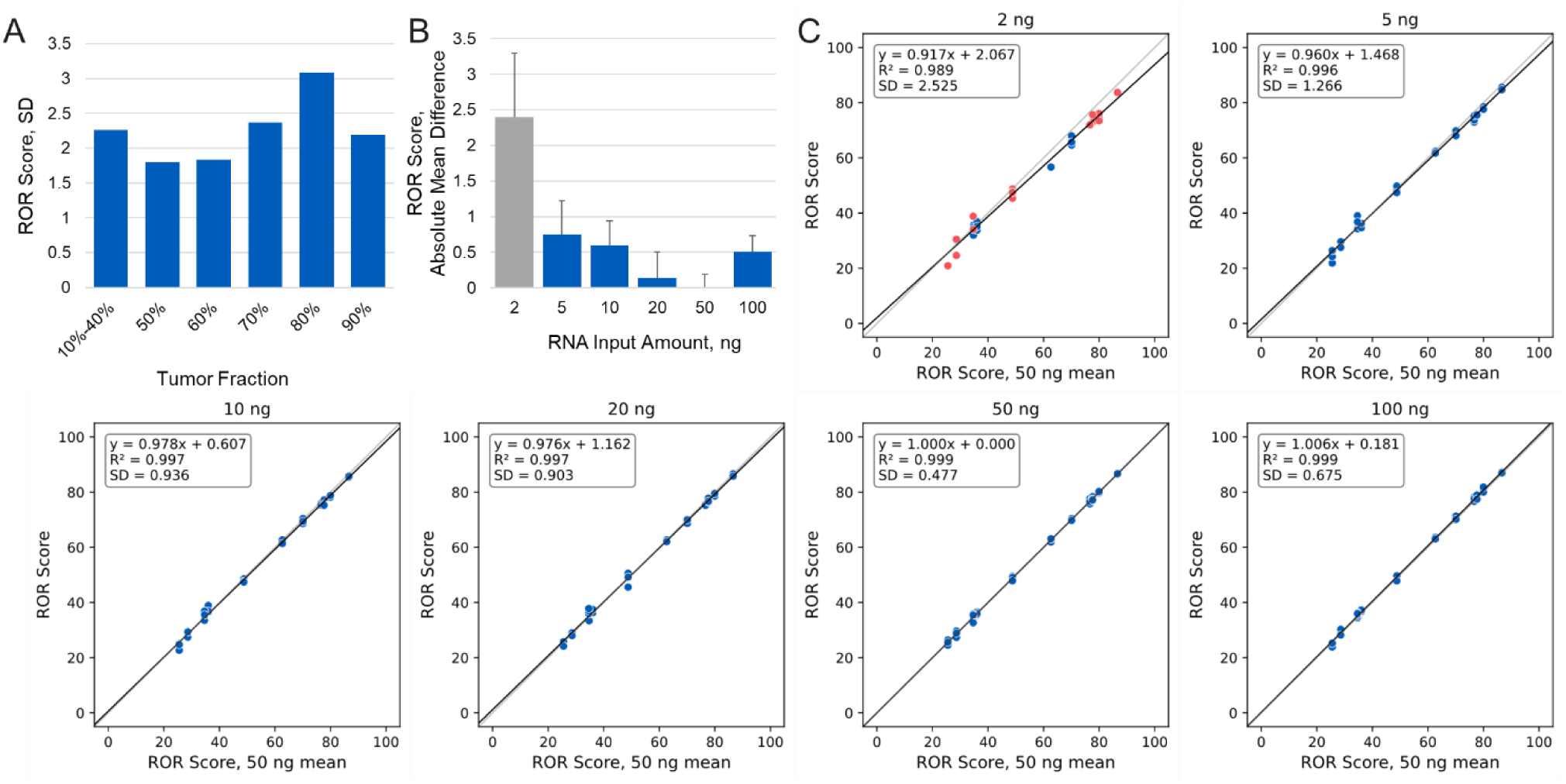
Analysis of NGS assay performance across different conditions. A) Analysis of tumor fraction in analytical validation cohort by pathologist assessment (n = 179). B) Assessment of lower limit of detection (LLOD) based on mean difference in ROR scores compared to standard 50 ng input. Conditions in gray failed to meet acceptance criteria (12 unique specimens, run in triplicate per condition). C) Regression of different RNA input amounts compared to mean ROR score calculated for each sample with 50ng input. Samples in red failed sequencing QC and were not analyzed for analytical performance but were included in the post-hoc regression analysis shown here.

The impact of excessive residual ethanol during RNA extraction or genomic DNA (gDNA) contamination was also evaluated as potential sources for contamination during extraction (**Figure 4B**). All interfering sample tests were run in triplicate with 50 ng RNA input. Two sets of 10 samples were chosen from either bridging or validation samples to be assessed for artificially increased ethanol or genomic DNA (gDNA) contamination. Ethanol was added to a final concentration of 10%, 20%, and 30% and all samples tested resulted in acceptable ROR score consistency (**Supplementary Figure 5**).

Two sources of gDNA contamination were separately tested as spiked-in contaminants. First, patient-derived genomic DNA was extracted using a sequential DNA/RNA extraction (MagMax, Thermo Fisher). Additionally, synthetic gDNA was prepared from sonicating DNA extracted from the NA12878 cell line. gDNA was spiked-in to extracted RNA samples at approximately 5% (2.5 ng), 10% (5 ng) and 20% (10 ng) prior to library preparation. Sheared genomic DNA (sDNA) resulted in 17 of 90 samples tested (18.9%) being flagged during NGS QC for contamination (**Supplementary Figure 6, Supplementary Table 6**). The samples for which NGS QC passed did not have significant variability in ROR scores. Additionally, genomic DNA directly extracted from FFPE samples did not result in sequencing QC flags after library preparation and was also observed to have acceptable variability in ROR scores.

### Retrospective analysis of long-term stored archival patient specimens

Finally, to demonstrate real-world validity on samples from archival material, the Prosigna NGS LDT was also run samples from the previously described Oslo1 study (19). Archival RNA (n = 109) was run in this study to evaluate performance of the LDT compared to previously reported Prosigna ROR scores from the nCounter system. The samples tested were long-term archival specimens, with RNA extracted following the Prosigna IVD instructions for use and stored at -80°C for more than ten years.

Testing the Prosigna NGS LDT on this real-world cohort of long-term archival specimens showed a high degree of concordance with nCounter system test results (**Figure 5**). The correlation between the previously run nCounter and NGS results was high (0.974, R^2^), and variance between the two tests was similar to the validation cohort (3.078, SD). We also observed high concordance between risk categories (95.4%, PWC; 0.944, weighted *κ*), and subtype classifications (94.5%, PWC; 0.927, unweighted *κ*) (**Supplementary Table 8**) (54). We also evaluated the performance of the Prosigna NGS LDT on RNA extracted from archival tissue blocks stored at RT (n = 28). Even though the blocks were very old (29 years old, median; 28 - 31range), none failed RNA extraction and only 4 failed NGS library preparation after PCR1. The Prosigna ROR scores calculated from RNA extracted from the archival blocks showed low variability (2.579, SD) and strong correlation (0.975, R^2^) with ROR scores calculated from the archival RNA (**Supplementary Figure 7**). Only one risk group classification change from intermediate to high risk was observed when comparing the Prosigna NGS test results calculated from archival RNA extractions or fresh extraction of RNA from archival FFPE blocks. No intrinsic subtype reclassifications were observed comparing archival FFPE blocks to archival RNA.

**Figure 5:**
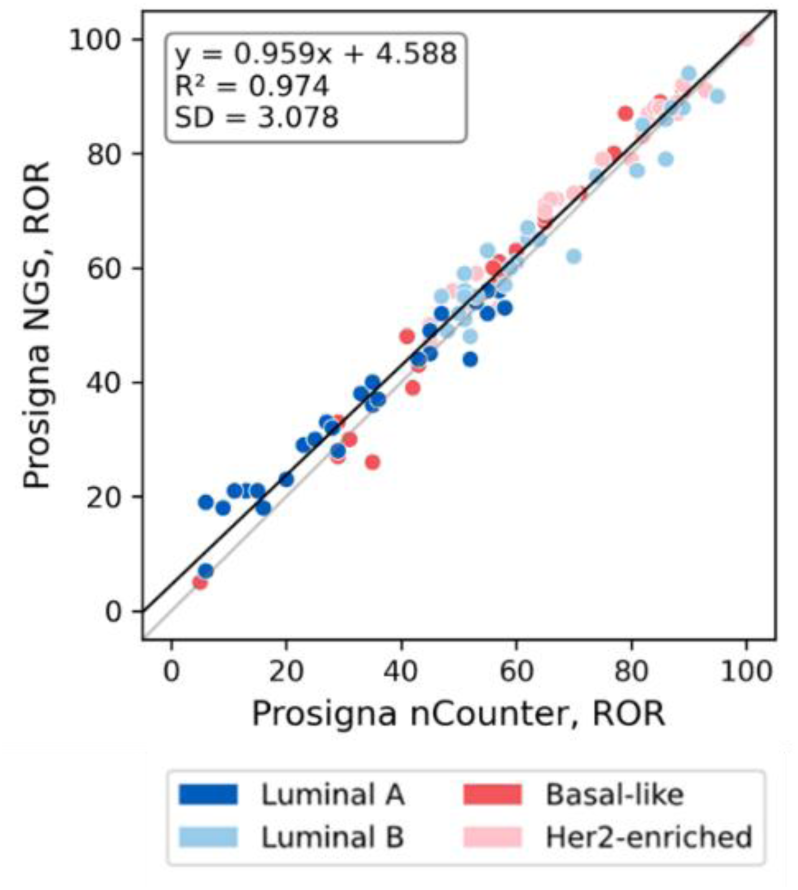
Comparison of performance of the Prosigna NGS LDT on archival RNA from Oslo1 clinical cohort (n = 109) extracted for the IVD Prosigna test for the nCounter system to original ROR scores from the nCounter system Prosigna test. Samples colored by intrinsic subtype called by NGS.

## Discussion

Breast cancer patients and their treating physicians have relied on GEP testing, including the Prosigna Breast Risk of Recurrence test, to help characterize factors driving tumor progression and response to intervention, for over two decades. This study validates leveraging whole transcriptome RNA sequencing at a central testing facility for determining the Prosigna ROR score and intrinsic subtype classification,. We validated performance on both SR and CNB FFPE tissue specimens to ensure the test met necessary requirements on samples routinely collected in both adjuvant and neoadjuvant settings. The strong correspondence between results generated on NGS and the nCounter system was expected, given the historical robustness of the key components of the Prosigna test, which spanned multiple expression profiling platforms. The intrinsic subtypes were initially derived from microarray studies (11, 12), PAM50 was later distilled to qPCR (1, 2), and ultimately validated analytically and clinically on the nCounter system (3–5, 13, 21, 39, 44).

The performance of the NGS LDT was robust in testing multiple cohorts of the intended test population, and across treatment conditions reflective of variables experienced with NGS testing. While the diagnostic input of 50 ng of RNA was validated here, we also show that the test is performant with minimal RNA input down to 5 ng and is robust to differences in tumor fraction of the tissue specimen. Throughout the study we show the robust Prosigna ROR score and intrinsic subtype calculation with as few as 3 FFPE slides, which is critical for studying CNB specimens which will have limited tissue availability. Additionally, the Prosigna NGS test was demonstrated to be robust to interfering substances that could potentially impact NGS performance, such as carry-over ethanol during RNA extraction and contaminating genomic DNA. Sheared exogenous DNA did create several flags during NGS testing, however these were appropriately caught by the sequencing pipeline and ultimately did not lead to high score variability when excluding samples. Further, the addition of patient-derived genomic DNA from the same FFPE sample did not impact test performance and is more reflective of real-world handling of the clinical specimens for this test.

Development and deployment of the Prosigna test as an LDT with central testing has several benefits for patients and clinicians. Clinicians are no longer required to have access to an nCounter system within their facilities, which removes the burden of both capital equipment purchase and specially trained laboratory staff. NGS testing has grown significantly in the last decade, and while the Prosigna test is not the first breast cancer prognostic GEP test to be validated on NGS (45), it is the first such breast cancer diagnostic GEP test that leverages whole transcriptome RNA sequencing. This provides the potential for the test to serve as a platform for ongoing translational research for future biomarkers. This study is strengthened through its use of multiple cohorts for both bridging and validation, with each cohort including diverse Prosigna ROR scores and intrinsic subtype classifications. As patients that are HR+ are classified as non-luminal subtypes in approximately 15% of cases (25, 26), this balancing ensures appropriate performance of the Prosigna NGS LDT for patients. Furthermore, the original development of PAM50 based intrinsic subtypes was derived to be performant in all breast cancer patients regardless of IHC subtype (1, 2, 39). We included a small fraction of HR-tumors in each cohort and balanced subtypes across the study to ensure appropriate performance, including in the clinical cohort (19). Additionally, that cohort included samples previously processed using IVD specifications for the Prosigna test to demonstrate that laboratory handling of tissues did not impact performance. Similarly, repeated runs on the same sample showed little variability attributable to operators, reagents or machine variability.

The scope of this study is limited to ensuring that the Prosigna NGS LDT can reliably report the Prosigna ROR score and intrinsic subtypes with high concordance to those reported from the nCounter system, and that the testing lab may be used later as a reference laboratory for further development. Additional studies to explore performance in other labs, and equivalence of NGS and nCounter results with distributed kits are still needed to ensure reliability in a distributed test format. Furthermore, while this study does evaluate two independent validation cohorts, we cannot at this time fully rule out that the wide range of processing variables involved with FFPE will not contribute to some level of assay variability not captured in this study. However, such impacts have been minimal on the nCounter test (55), and is unlikely given the strong performance observed on decades-old samples from the Oslo1 study. Similarly, while different RNA extraction methods may result in noticeable differences in DV200 levels, whole exome capture techniques have not been reported to be significantly influenced by variation in RNA degradation in FFPE tissues (48, 56). Finally, this study is limited to evaluating the test scores and constituent components of the Prosigna test. Previous studies have tested the performance of the reagents used as part of this platform in our labs and others (46, 48, 52, 53), and subsequent studies should be conducted focused on the translational and clinical utility of the broader platform.

## Conclusions

The development and validation of the Prosigna Breast Risk of Recurrence test on NGS as an LDT modernizes the underlying technology for clinical determination of the risk of recurrence and intrinsic subtype for early-stage breast cancer patients. We observed high concordance and low variability across all studies needed to demonstrate that this LDT is equivalent to the previously validated test on the nCounter system. This study shows that the Prosigna test can confidently support patients who face critical decisions about their care and treatment after their initial diagnosis.

## Data Availability

The datasets analyzed during the current study are not publicly available due to proprietary commercial development but are available from the corresponding author on reasonable request.

## Declarations

### Ethics approval and consent to participate

The study was approved by the Norwegian Regional Committees for Medical and Health Research Ethics, REC South East (reference 2015/2453). Written consent was obtained from all patients.

---

**Supplementary Figure 1:**
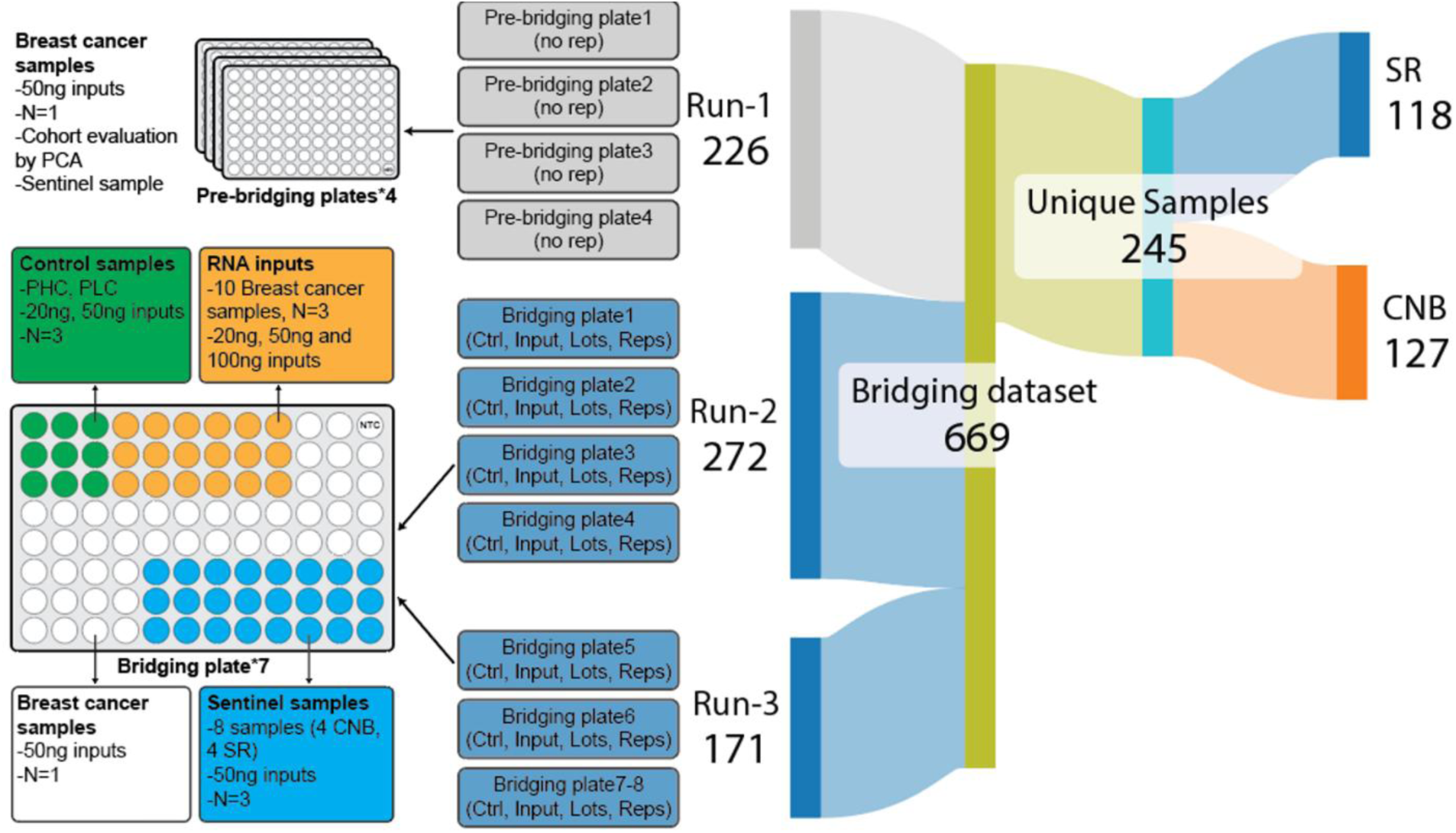
Sample organization and plates tested during NGS LDT bridging study.

**Supplementary Figure 2:**
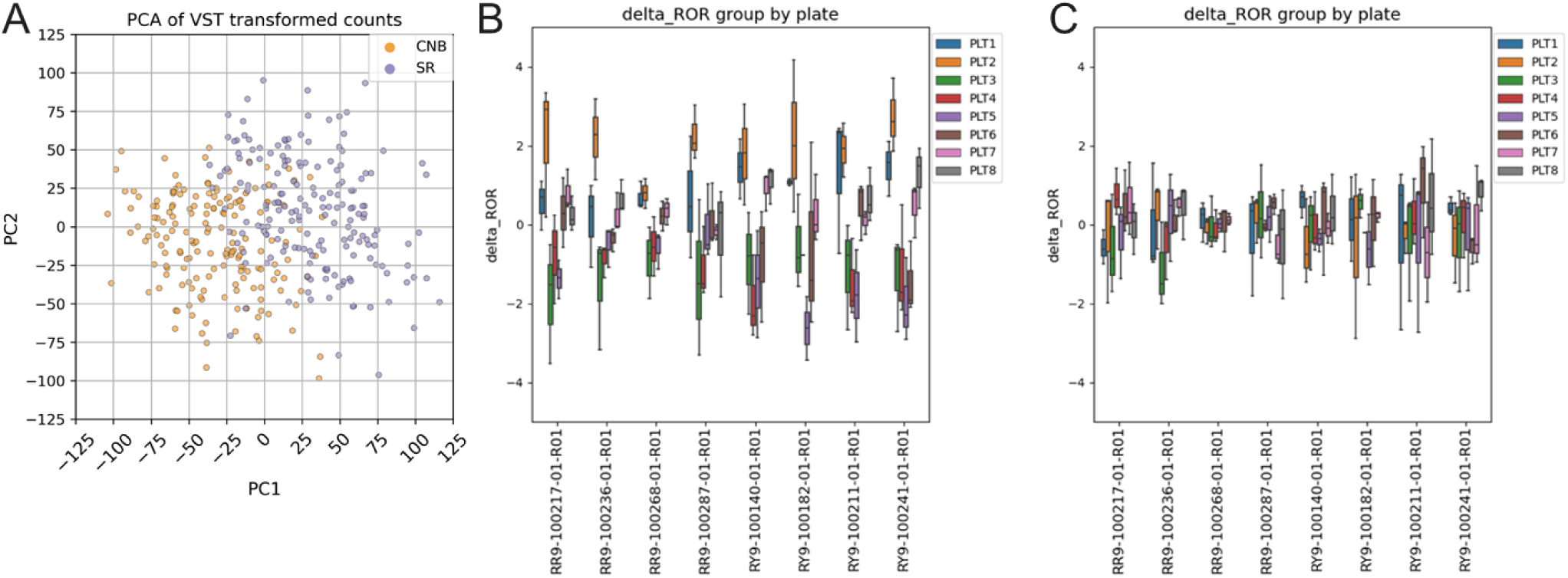
PCA Analysis of SR and CNB samples from bridging study. A) Analysis of VST normalized expression of all unique samples from bridging study. Samples colored by specimen type. B-C) Evaluation of batch correction on differences in Prosigna ROR score showing plotting differences in ROR scores between platforms for sentinel samples (x-axis) grouped by sequencing plate either B) before or C) after batch correction.

**Supplementary Figure 3:**
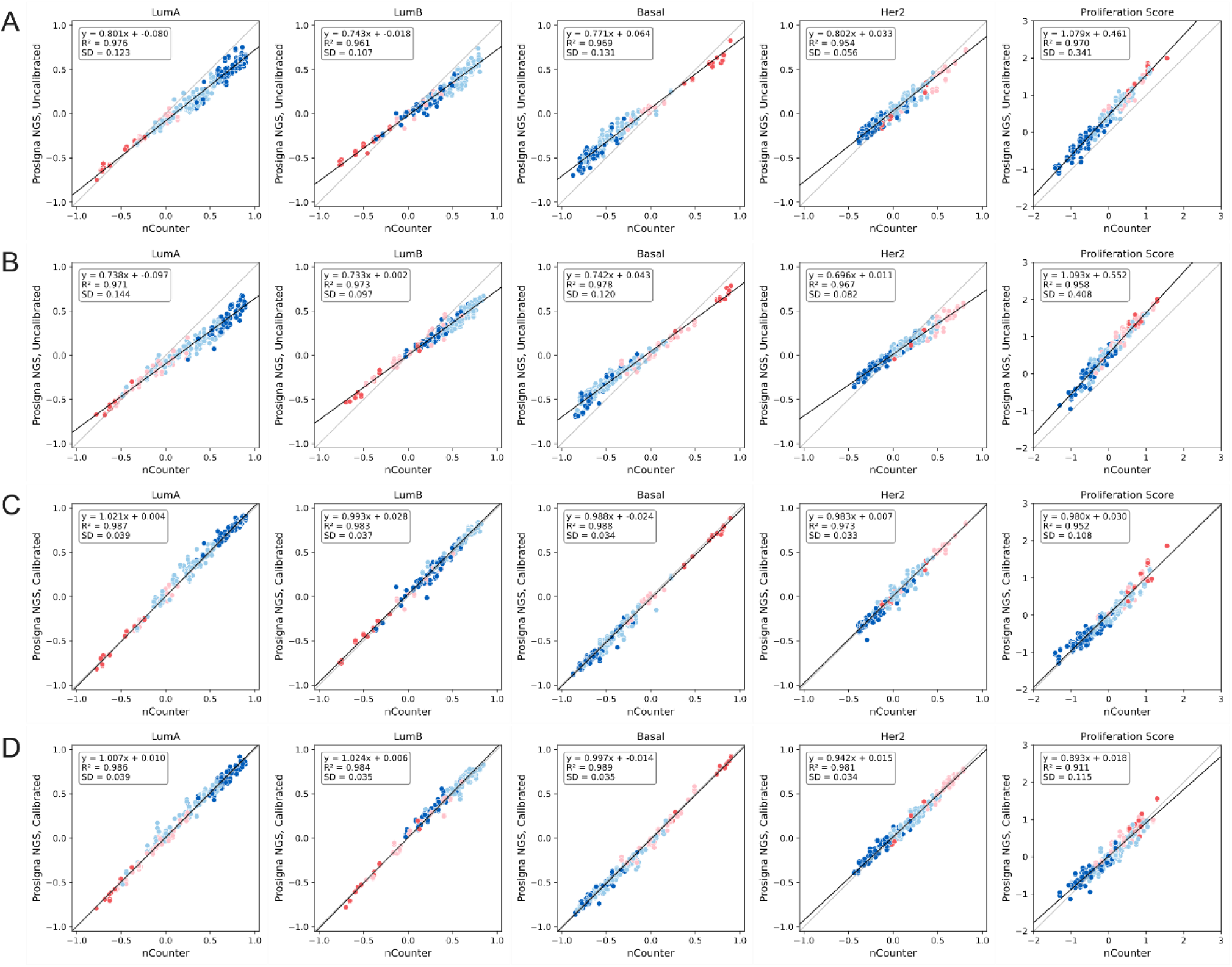
Component scores for Prosigna ROR and intrinsic subtype classification for bridging study (n = 245) samples before (A, B) or after (C, D) calibration for SR (A, C), and CNB (B, D) samples. Intrinsic subtype as classified by NGS is shown. Linear regression line shown in black.

**Supplementary Figure 4:**
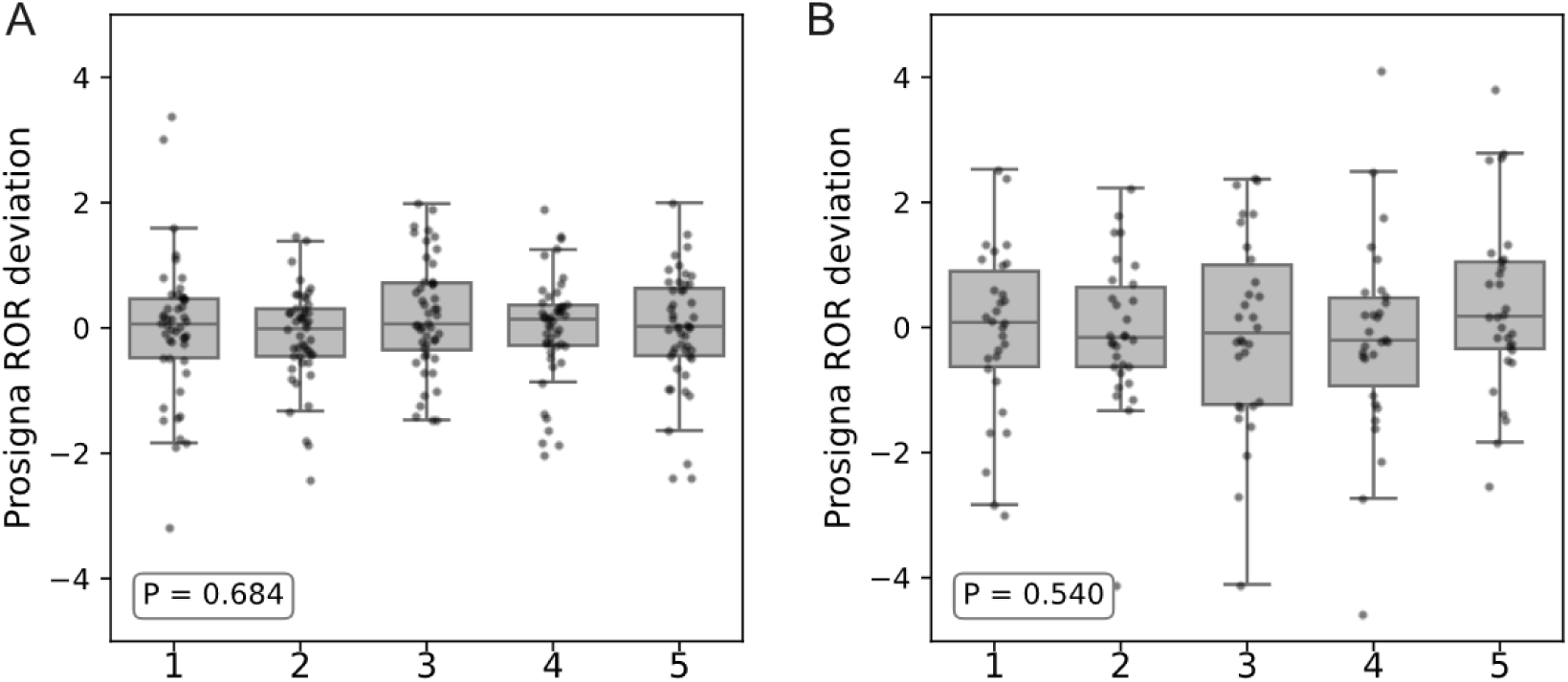
Evaluation of analytical precision among sequencing runs comparing deviation in Prosigna ROR scores. Each group represents a sequencing plate. Precision shown from A) library preparation or B) RNA extraction and library preparation. ANOVA P-value is shown.

**Supplementary Figure 5:**
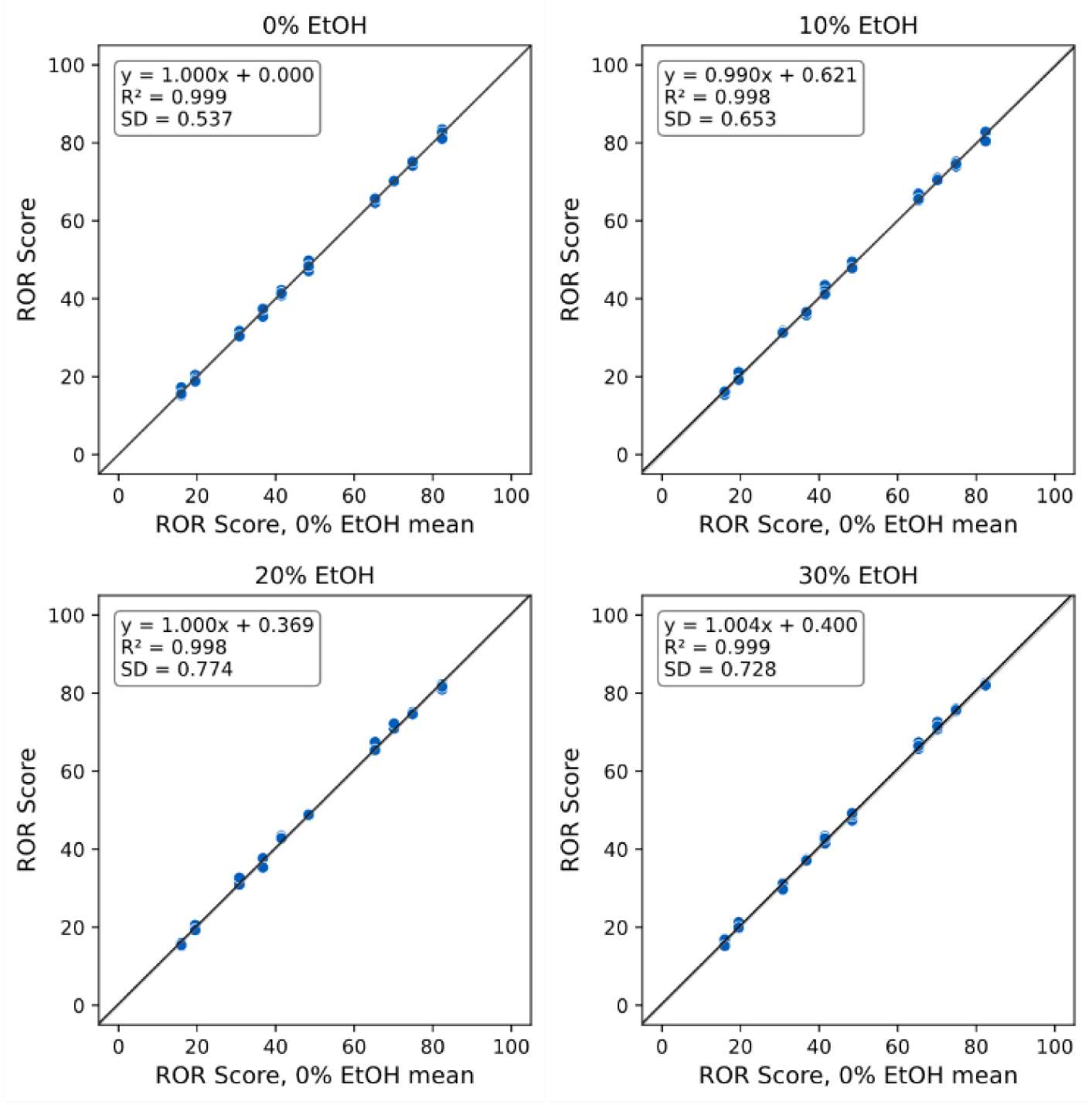
Impact of ethanol contamination during interfering substances test (10 unique samples, run in triplicate per condition). Prosigna ROR scores from samples with various concentrations of ethanol added to extracted RNA samples compared to no contamination control mean.

**Supplementary Figure 6:**
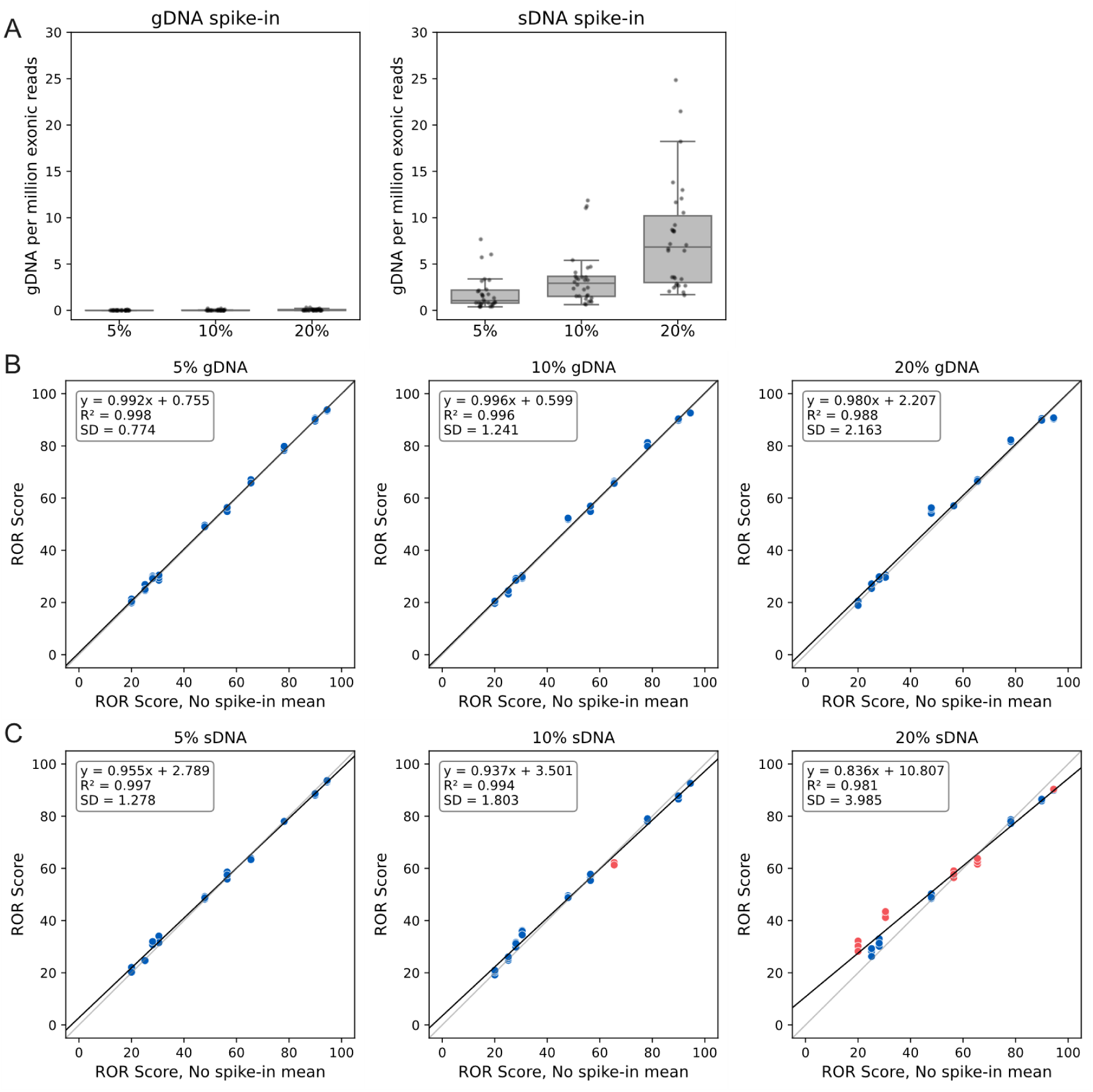
Impact of genomic DNA contamination during interfering substances test (10 unique samples, run in triplicate per condition). A) Incorporation of genomic DNA detected in sequencing QC. Threshold for acceptability set at < 7.68 gDNA per million exonic reads. Samples separated based on source of genomic DNA for patient derived (left) and synthetic cell line sheered DNA (right). B-C) ROR scores for samples from patient-derived (B, gDNA), and synthetic cell line sheered DNA (C, sDNA). Samples in red failed sequencing QC and were not analyzed for analytical performance but were included in the post-hoc regression analysis shown here.

**Supplementary Figure 7:**
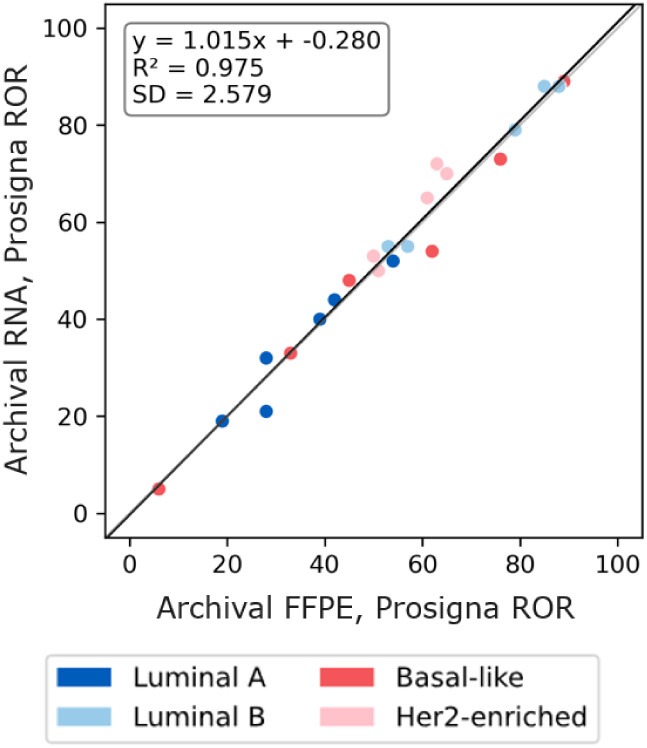
Comparison of Oslo1 clinical validation cohort with the Prosigna NGS LDT on archival RNA extraction stored at -80C orRNA extraction from archival FFPE blocks stored at RT.

**Supplementary Table 1:** Results table from analytical validation testing.

| Study | # Samples / # data points | Acceptance Criteria | Results |
| --- | --- | --- | --- |
| **Accuracy** | 187 / 187<br>(104 SR and 83 CNB) | Deviation in ROR from nCounter system test $SD \leq 5$. | SR samples=2.488;<br>CNB samples=2.588 |
| | | intrinsic subtype concordance $\geq 85\%$. | SR samples=92.31%;<br>CNB samples=92.77% |
| | | Risk group concordance for node negative and node positive samples with thresholds at 40 and 60: Major reclass rate $\leq 5\%$. | SR samples= 0%;<br>CNB samples= 0% |
| | | Risk group concordance for node positive samples with threshold of 40 only (prognostic): $\geq 80\%$ | SR samples= 97.67%;<br>CNB samples=95.45% |
| **Precision of RNA Extraction** | 10 / 150 | Repeatability: ROR $SD \leq 4.3$<br>Reproducibility ROR $SD \leq 4.3$ across conditions. | Repeatability $SD=1.462$;<br>Reproducibility=1.462 |
| | | Repeatability risk category: PWC $\geq 80\%$<br>Repeatability subtype: PWC $\geq 80\%$<br>Reproducibility risk category: PWC $\geq 80\%$<br>Reproducibility subtype: PWC $\geq 80\%$ | Repeatability risk category PWC=95.1%;<br>Repeatability subtype PWC=100.0%;<br>Reproducibility risk category PWC=95.2%;<br>Reproducibility subtype PWC=100.0% |
| **Precision of library preparation from RNA** | 16 / 240 | Repeatability: ROR $SD \leq 4.3$<br>Reproducibility ROR $SD \leq 4.3$ across conditions. | Repeatability $SD=0.951$;<br>Reproducibility=0.951 |
| | | Repeatability risk category: PWC $\geq 80\%$<br>Repeatability subtype: PWC $\geq 80\%$<br>Reproducibility risk category: PWC $\geq 80\%$<br>Reproducibility subtype: PWC $\geq 80\%$ | Repeatability risk category PWC=100.0%;<br>Repeatability subtype PWC=96.7%;<br>Reproducibility risk category PWC= 100.0%;<br>Reproducibility subtype PWC=97.4% |
| **Extraction Method Equivalency** | 10 / 60 | $SD$ of ROR $\leq 4.3$ across conditions. | $SD= 0.754$ |
| **Lower Limit of Tumor Fraction (LLoTF)** | 178 / 178 | Deviation in ROR from nCounter system test: $SD \leq 5$ for any tumor fraction quantile. | Minimum Tumor Content 10% ($SD=2.27$) |
| **Analytical Sensitivity, input range (LLoD)** | 12 / 193 | The test condition will be considered equivalent to the nominal level if the 90% confidence interval is entirely contained within the $-3$ to $3$ ROR range. | 5 ng minimum input (90% CI: [-1.22, -0.29]) |
| | | Deviation in ROR from standard condition $SD \leq 5$. | 2 ng minimum input ($SD=2.33$) |
| **Interfering substances of genomic DNA in RNA samples** | 10 / 193 | The test condition will be considered equivalent to the nominal level if the 90% confidence interval is entirely contained within the $-3$ to $3$ ROR range. | Up to 20% gDNA content (Sample level gDNA 90% CI=[-0.14,0.94]; sheared gDNA 90% CI=[-1.16, 1.45]) |
| | | Deviation in ROR from standard condition $SD \leq 5$. | Up to 20% gDNA content (Sample level gDNA $SD=2.16$; sheared gDNA $SD=2.12$) |
| **Interfering substances of ethanol in RNA samples** | 10 / 120 | The test condition will be considered equivalent to the nominal level if the 90% confidence interval is entirely contained within the $-3$ to $3$ ROR range. | Up to 30% EtOH spike in (90% CI= [0.36, 0.87]) |
| | | Deviation in ROR from standard condition $SD \leq 5$. | Up to 30% EtOH spike in ($SD=0.73$) |
| Sequencing Reproducibility and flow cell type evaluation | 96 samples, 10B flow cell | SD of ROR 4.3 across conditions. | SD=0.73 |
| | | PWC $\geq 80\%$ | PWC=96.8% |
|  | 36 samples, 1.5B flow cell | SD of ROR 4.3 across conditions. | SD=0.66 |
| | | PWC $\geq 80\%$ | PWC=97.1% |
|  | 96 samples, 1.5B flow cell | SD of ROR 4.3 across conditions. | SD=0.938 |
| | | PWC $\geq 80\%$ | PWC=97.8% |

**Supplementary Table 2:** TOST analysis for the Prosigna ROR score from matched NGS LDT and nCounter system test IVD kit. Equivalence bounds of ±3 ROR units were used. Equivalence is concluded when both one-sided tests are significant (i.e., when this maximum p-value is less than the significance level).

|  | Mean difference | 90%CI | TOST Lower p-val | TOST Upper p-val |
| --- | --- | --- | --- | --- |
| **SR only** | 1.377 | [0.848, 1.907] | 2.77E-25 | 8.25E-07 |
| **CNB only** | -0.218 | [-0.882, 0.445] | 3.60E-10 | 2.54E-12 |
| **SR +CNB** | 0.669 | [0.245, 1.093] | 4.57E-32 | 7.76E-17 |

**Supplementary Table 3:**
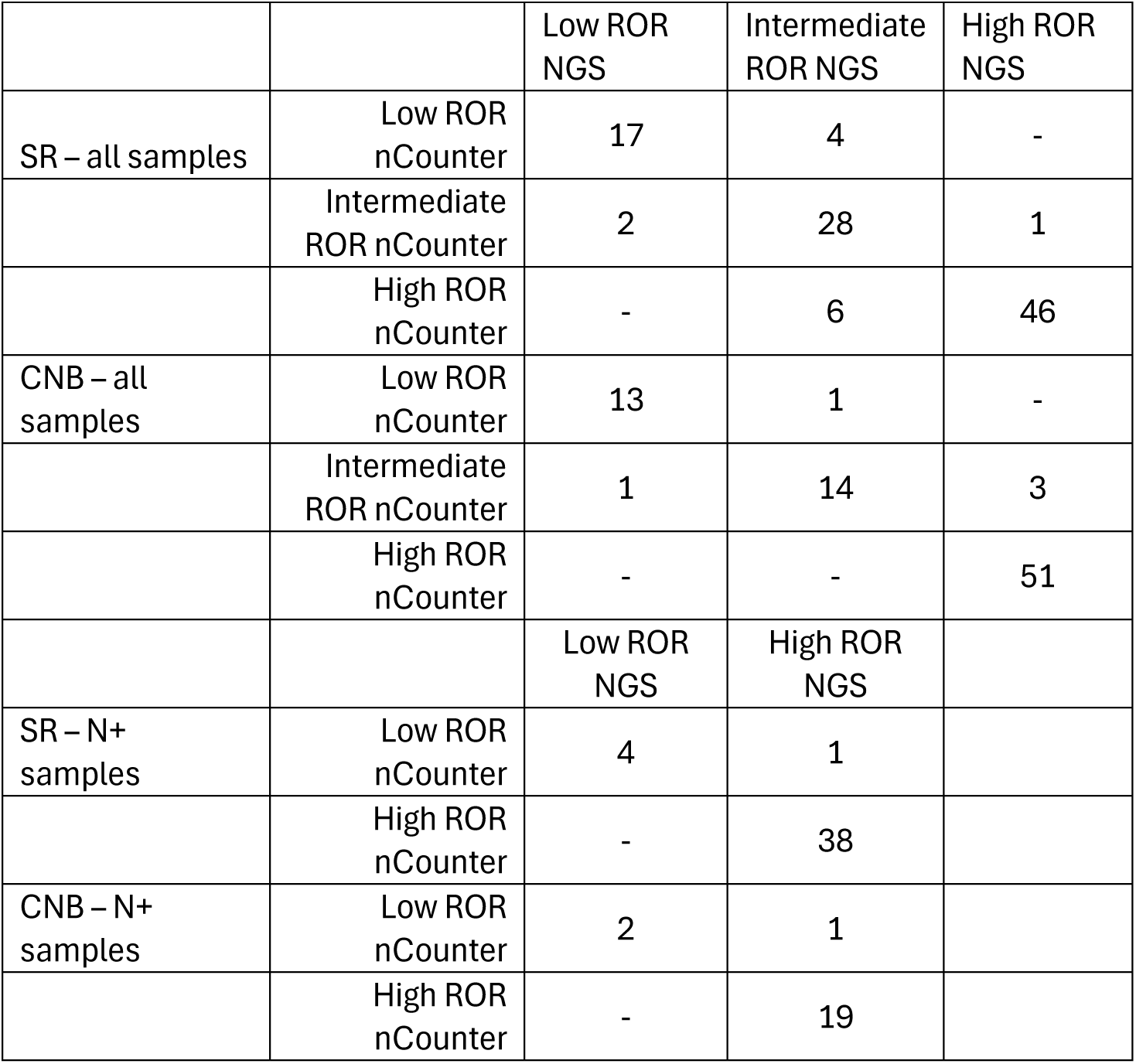
Confusion matrices for ROR risk categories in all samples by specimen type or for major risk categories in node positive patients for the analytical validation study.

|  |  | Low ROR<br>NGS | Intermediate<br>ROR NGS | High ROR<br>NGS |
| --- | --- | --- | --- | --- |
| SR – all samples | Low ROR<br>nCounter | 17 | 4 | - |
|  | Intermediate<br>ROR nCounter | 2 | 28 | 1 |
|  | High ROR<br>nCounter | - | 6 | 46 |
| CNB – all<br>samples | Low ROR<br>nCounter | 13 | 1 | - |
|  | Intermediate<br>ROR nCounter | 1 | 14 | 3 |
|  | High ROR<br>nCounter | - | - | 51 |
|  |  | Low ROR<br>NGS | High ROR<br>NGS |  |
| SR – N+<br>samples | Low ROR<br>nCounter | 4 | 1 |  |
|  | High ROR<br>nCounter | - | 38 |  |
| CNB – N+<br>samples | Low ROR<br>nCounter | 2 | 1 |  |
|  | High ROR<br>nCounter | - | 19 |  |

**Supplementary Table 4:** Confusion matrices for IS classification for all samples by specimen type for analytical validation study.

|  |  | Luminal A<br>NGS | Luminal B<br>NGS | Basal-like<br>NGS | Her2-enriched<br>NGS |
| --- | --- | --- | --- | --- | --- |
| SR samples | Luminal A<br>nCounter | 37 | 3 | - | - |
|  | Luminal B<br>nCounter | 3 | 45 | - | - |
|  | Basal-like<br>nCounter | 1 | - | 5 | - |
|  | Her2-enriched<br>nCounter | - | 1 | - | 9 |
| CNB samples | Luminal A<br>nCounter | 21 | 2 | - | - |
|  | Luminal B<br>nCounter | 1 | 39 | - | - |
|  | Basal-like<br>nCounter | - | 1 | 6 | 1 |
|  | Her2-enriched<br>nCounter | - | 1 | - | 11 |

**Supplementary Table 5 -.**
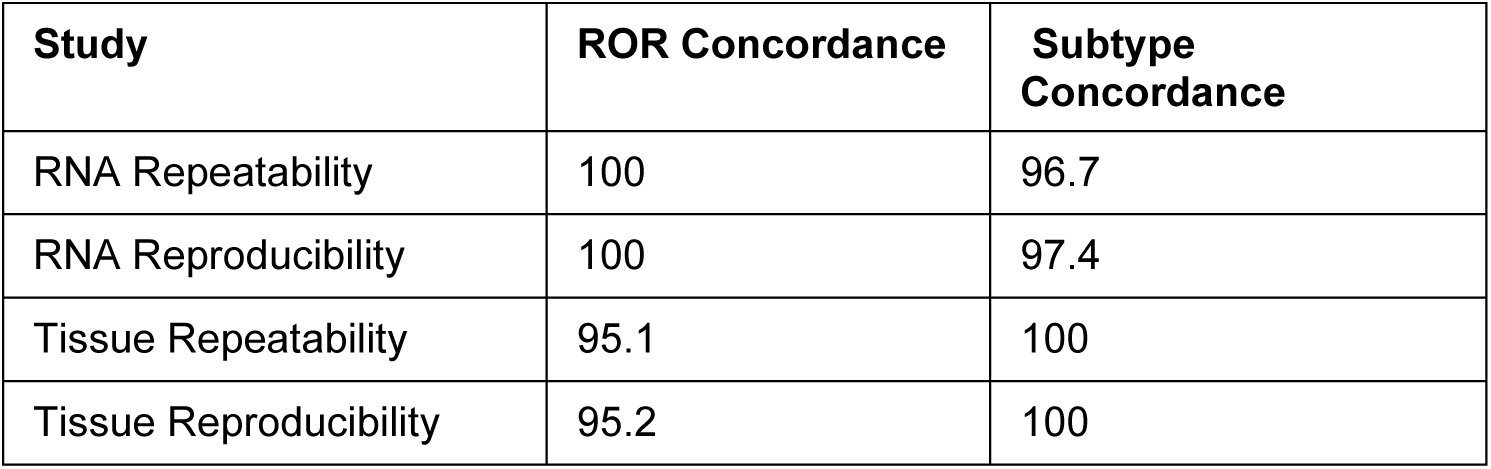
Repeatability (inter-run) and reproducibility (intra-run) of ROR scores and intrinsic subtypes from repeated sequencing from the same RNA pool or repeated extraction from the same tissue block.

| **Study** | **ROR Concordance** | **Subtype Concordance** |
| --- | --- | --- |
| RNA Repeatability | 100 | 96.7 |
| RNA Reproducibility | 100 | 97.4 |
| Tissue Repeatability | 95.1 | 100 |
| Tissue Reproducibility | 95.2 | 100 |

**Supplementary Table 6:** LLOD and interfering substances testing details.

| **Evaluation** | **QC**<br>**Flagged**<br>**Samples** | **Input**<br>**Source** | **Condition** | **N** | **Mean**<br>**difference** | **CI**<br>**lower**<br>**limit** | **CI upper**<br>**limit** | **Passed**<br>**Samples**<br>**(#)** | **Mean difference**<br>**(Confidence**<br>**Interval)** |
| --- | --- | --- | --- | --- | --- | --- | --- | --- | --- |
| LLOD | Included | N/A | 2ng | 27 | -2.4 | -3.3 | -1.5 | 13 | -2.4 (-3.3,-1.5) |
| LLOD | Included | N/A | 5ng | 36 | -0.75 | -1.22 | -0.29 | 36 | -0.75 (-1.22,-0.29) |
| LLOD | Included | N/A | 10ng | 36 | -0.6 | -0.94 | -0.27 | 36 | -0.6 (-0.94,-0.27) |
| LLOD | Included | N/A | 20ng | 36 | -0.13 | -0.5 | 0.23 | 36 | -0.13 (-0.5,0.23) |
| LLOD | Included | N/A | 50ng | 36 | 0 | -0.19 | 0.19 | 36 | 0 (-0.19,0.19) |
| LLOD | Included | N/A | 100ng | 36 | 0.5 | 0.27 | 0.73 | 36 | 0.5 (0.27,0.73) |
| LLOD | Excluded | N/A | 2ng | 13 | -2.2 | -3.46 | -0.93 | 13 | -2.2 (-3.46,-0.93) |
| LLOD | Excluded | N/A | 5ng | 36 | -0.75 | -1.22 | -0.29 | 36 | -0.75 (-1.22,-0.29) |
| LLOD | Excluded | N/A | 10ng | 36 | -0.6 | -0.94 | -0.27 | 36 | -0.6 (-0.94,-0.27) |
| LLOD | Excluded | N/A | 20ng | 36 | -0.13 | -0.5 | 0.23 | 36 | -0.13 (-0.5,0.23) |
| LLOD | Excluded | N/A | 50ng | 36 | 0 | -0.19 | 0.19 | 36 | 0 (-0.19,0.19) |
| LLOD | Excluded | N/A | 100ng | 36 | 0.5 | 0.27 | 0.73 | 36 | 0.5 (0.27,0.73) |
| gDNA | Included | gDNA | 0pct | 30 | 0 | -0.22 | 0.22 | 30 | 0 (-0.22,0.22) |
| gDNA | Included | gDNA | 5pct | 30 | 0.32 | -0.01 | 0.65 | 30 | 0.32 (-0.01,0.65) |
| gDNA | Included | gDNA | 10pct | 30 | 0.4 | -0.14 | 0.94 | 30 | 0.4 (-0.14,0.94) |
| gDNA | Included | gDNA | 20pct | 30 | 1.14 | 0.24 | 2.03 | 30 | 1.14 (0.24,2.03) |
| gDNA | Included | sDNA | 5pct | 30 | 0.38 | -0.18 | 0.94 | 29 | 0.38 (-0.18,0.94) |
| gDNA | Included | sDNA | 10pct | 30 | 0.14 | -0.66 | 0.94 | 27 | 0.14 (-0.66,0.94) |
| gDNA | Included | sDNA | 20pct | 30 | 2 | 0.34 | 3.67 | 17 | 2 (0.34,3.67) |
| gDNA | Excluded | sDNA | 5pct | 29 | 0.45 | -0.12 | 1.01 | 29 | 0.45 (-0.12,1.01) |
| gDNA | Excluded | sDNA | 10pct | 27 | 0.55 | -0.23 | 1.34 | 27 | 0.55 (-0.23,1.34) |
| gDNA | Excluded | sDNA | 20pct | 17 | 0.15 | -1.16 | 1.45 | 17 | 0.15 (-1.16,1.45) |
| EtOH | N/A | N/A | 0pct | 30 | 0 | -0.24 | 0.24 | 30 | 0 (-0.24,0.24) |
| EtOH | N/A | N/A | 10pct | 30 | 0.15 | -0.14 | 0.43 | 30 | 0.15 (-0.14,0.43) |
| EtOH | N/A | N/A | 20pct | 30 | 0.35 | 0.02 | 0.68 | 30 | 0.35 (0.02,0.68) |
| EtOH | N/A | N/A | 30pct | 30 | 0.61 | 0.35 | 0.87 | 30 | 0.61 (0.35,0.87) |

**Supplementary Table 7:** RNA extraction yields from 10 specimens extracting from either five 5 µm sections or three 10 µm section.

| RNA Concentration (ng/μL) |  |
| --- | --- |
| Five 5-μm sections | Three 10-μm sections |
| 88.0 | 84.1 |
| 389.3 | 436.4 |
| 17.1 | 26.0 |
| 99.0 | 61.1 |
| 68.3 | 89.1 |
| 343.0 | 289.3 |
| 244.9 | 207.2 |
| 86.1 | 79.6 |
| 55.7 | 72.9 |
| 47.0 | 73.5 |

**Supplementary Table 8:** Confusion matrices for Prosigna ROR risk categories and intrinsic subtype classification for analytical the Oslo1 clinical validation study of archival RNA specimens.

|  | Low ROR<br>NGS | Intermediate<br>ROR NGS | High ROR<br>NGS |  |
| --- | --- | --- | --- | --- |
| Low ROR nCounter | 22 | 0 | 0 |  |
| Intermediate ROR nCounter | 1 | 36 | 4 |  |
| High ROR nCounter | 0 | 0 | 46 |  |
|  | Luminal A<br>NGS | Luminal B<br>NGS | Basal-like<br>NGS | Her2-enriched<br>NGS |
| Luminal A nCounter | 24 | 3 | 0 | 0 |
| Luminal B nCounter | 1 | 26 | 0 | 0 |
| Basal-like nCounter | 0 | 0 | 26 | 1 |
| Her2-enriched nCounter | 0 | 1 | 0 | 27 |

